# Biological sex affects gene expression and functional variation across the human genome

**DOI:** 10.1101/2024.09.03.24313025

**Authors:** Angela G. Jones, Trisha Dalapati, Guinevere G. Connelly, Liuyang Wang, Benjamin H. Schott, Adrianna K. San Roman, Dennis C. Ko

## Abstract

Humans display sexual dimorphism across many traits, but little is known about underlying genetic mechanisms and impacts on disease. We utilized single-cell RNA-seq of 480 lymphoblastoid cell lines to identify 1200 genes with significantly sex-biased expression. While reproducibility was highest among LCL datasets, 71% were found to be sex-biased in at least one GTEx tissue, with a core dataset of 21 genes displaying sex-biased expression across all datasets and tissues examined. While 7.7% of sex-biased genes can be directly explained by differences in the number of sex chromosomes, most sex-biased genes (79%) are targets of transcription factors that display sex-biased expression. FOSL1, ZNF730, ZFX, and ZNF726 appear to make the largest contribution to this based on machine learning and linear modeling approaches, and all four of these transcription factors are regulated by the number of X chromosomes. Further, by testing the difference in slopes of conditionally independent expression quantitative trait loci (eQTL) identified in each sex separately, we identified 2,390 sex-biased eQTL (sb-eQTL) across the genome. While evidence of replication in an independent dataset was modest, permutation analysis demonstrated that sb-eQTL identified using real sex was more likely to have concordant direction of effect. These sb-eQTL are enriched in over 100 GWAS phenotypes, including many loci associated with female-biased autoimmune diseases such as multiple sclerosis. Our results demonstrate widespread genetic impacts on sexual dimorphism and identify possible mechanisms and clinical targets for sex differences in diverse diseases.

---

Humans display sexual dimorphism across a variety of traits. Anthropometric and physiological traits (e.g., body height, waist-to-hip ratio, etc.) are obvious examples of differences between males and females, but sex differences are also evident in the incidence, prevalence, severity, and treatment response in human disease. For example, females are more likely to develop autoimmune disease^1,2^ but show greater resistance to many infectious agents than males^3^. Sex disparities are also apparent across cancers, with kidney^4^, liver^5^, skin^6^, and laryngeal ^7^ cancers displaying a male bias, while breast and thyroid cancers ^8^ are more common in females. Phenotypic sex differences have been attributed to various factors including sex-specific hormone activity, sex chromosome complement, sex and gender influences on behavior, and complex differences in environment. However, despite these widespread differences, little is known about the mechanisms and biological functions that underlie sexual dimorphism in humans.

Previous studies have shown that gene expression differs by sex throughout the genome^9,10^, and several genome-wide association studies (GWAS) have identified genetic variants that show sex-specific effects in human disease^11–14^. However, while previous studies have identified genes with sex-biased expression (sb-Genes)^10,15,16^, how well sb-Genes replicate across diverse datasets and the underlying molecular mechanisms of why individual genes display sex-biased expression are not well characterized. Further, recent attempts to link gene expression differences to single genetic variants through sex-biased expression quantitative trait loci (sb-eQTL) discovery have reported few significant associations, with little replication across independent datasets^10,17–19^, leading to uncertainty in whether sb-eQTL contributed meaningfully to sex-biased expression^10^. The lack of sb-eQTL previously discovered and replicated may be due to relatively small differences in the genetic control of gene expression by sex, low power in previous studies, or tissue-type specificity of genetic effects.

Here, we performed single-cell RNA-seq of 240 male and 240 female lymphoblastoid cell lines (LCLs) to identify transcriptomic sex differences and sb-eQTL across the genome. We identified 1,200 sb-Genes, replicated these in multiple datasets across tissues, and categorized their effects by underlying mechanism. Although a two-step method identified 3,679 sb-eQTL, we found that sb-eQTL are not a major factor in sex-biased expression, although likely still contribute to sex-biased traits at individual loci. In contrast, we were able to assign ∼97% of all sb-Genes to mechanisms involving sex chromosome copy number or transcription factor expression, revealing a molecular basis for sex-biased gene expression throughout the genome.

## Sex-biased gene expression is reproducible and a subset is generalizable across tissues

Previous work in identifying sex-biased gene expression has identified thousands of genes displaying sex-biased expression (sb-Genes)^10,16^. The GTEx study examined conservation of sb-Genes across tissues and found largely tissue-specific effects, although a fraction of sb-Genes enriched for XCI-escapees were consistent across tissues. However, it is important to determine how well the sb-Genes identified in this ground-breaking work replicate across different individuals, particularly using a dataset with diverse genetic ancestry. Therefore, we sought to identify genes with sex-biased expression (sb-Genes) in a single cell type that would allow for testing replication in multiple datasets.

We previously developed a rapid and scalable pooled scRNA-seq method to uncover transcriptomic differences in a diverse cohort of 96 LCLs called scHi-HOST (single-cell high-throughput human in vitro susceptibility testing)^20^. We expanded this method to 480 LCLs in twelve worldwide populations (**Fig. 1A**). Through integration of phased whole genome sequencing, we are able to deconvolute pooled transcriptomics to identify gene expression from each individual LCL, allowing for rapid screening of hundreds of cell lines. Importantly for this study, scHi-HOST was conducted with equal numbers of male and female LCLs, as defined by sex chromosome complement. Through this controlled method, we can isolate sex-specific transcriptomic signatures due to genetic differences without the impact of differential hormones and environmental conditions. In total, we sequenced 196,338 single cells that mapped to a single individual, with a mean of 409 cells per individual (**Table S1; Fig. S1**). As expected, we detected highly significant differences of X and Y chromosomal genes that matched the reported sex of these LCLs (**Fig. S1C**) but also observed significant changes in autosomal genes. In total, we discovered 1,200 genes with significant differential expression by sex (sb-Genes, FDR<0.05; **Fig. 1B**; **Table S2**). Also as expected, the effect size of sex-bias was largest for Y chromosomal genes (mean |log2FC|=6.59, 3.56-7.96), followed by X chromosomal genes (mean |log2FC|=0.34, 0.04-7.22), with autosomal genes showing the smallest effect (mean |log2FC|=0.02, 0.02-1.88; **Fig. 1C**). KEGG enrichment revealed sb-Gene functional enrichment in pathways involving immune responses and basic cellular functions (**Fig. S1**).

**Figure 1.**
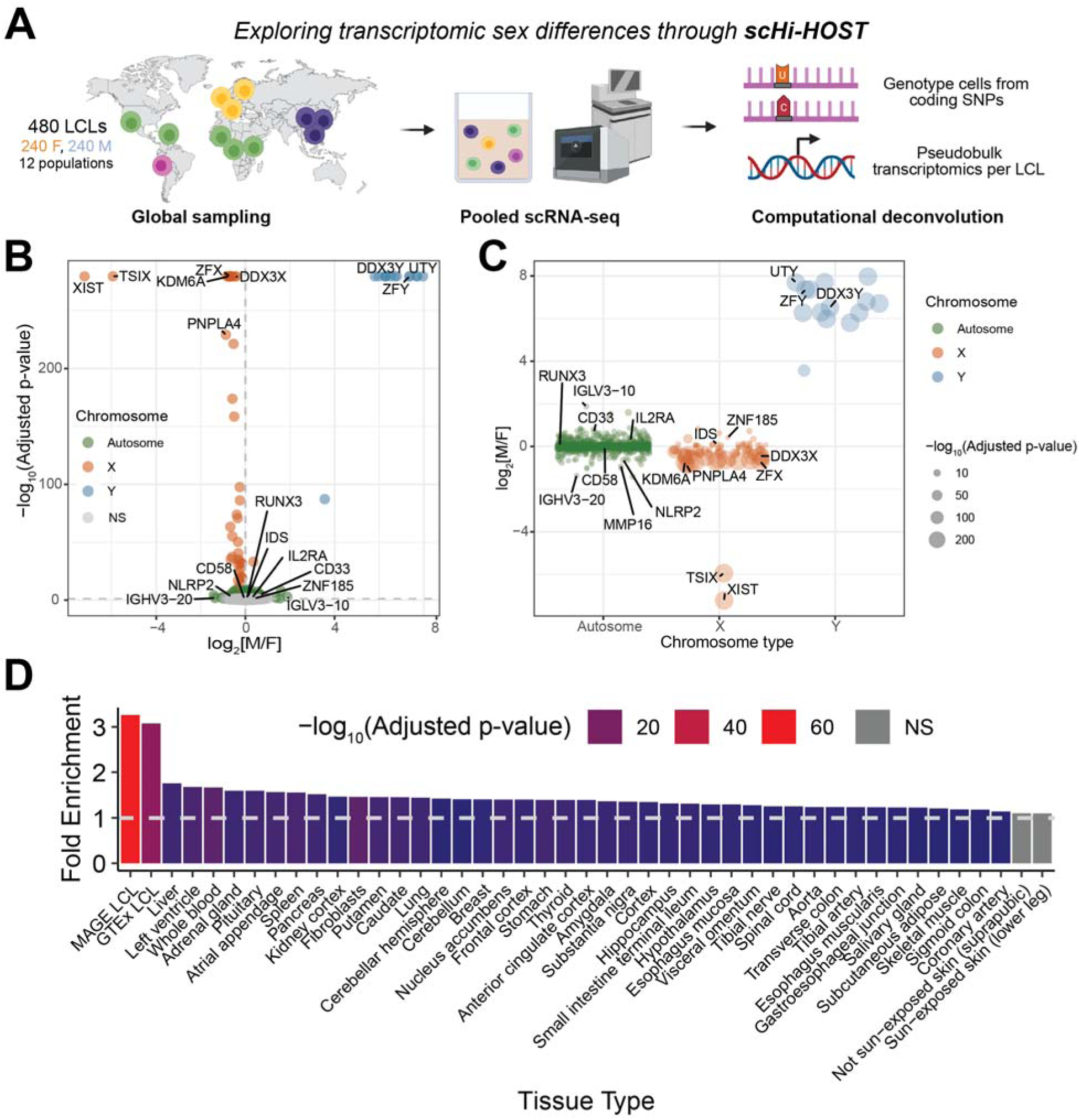
Sex-biased expression in LCLs is replicable and partly conserved across tissues. (**A**) Schematic of scHi-HOST pipeline. (**B**) Volcano plot of differential expression by sex colored by chromosomal location (FDR<0.05). (**C**) Sex-biased genes (FDR<0.05) displayed by chromosomal location. (**D**) Enrichment of sex-biased genes (FDR<0.05) across tissue types. Fisher’s Exact Tests were conducted for each tissue type.

We assessed the level of replication of sb-Genes in an independent LCL dataset of comparable power (MAGE^21^) and across human tissues with the GTEx dataset^1^ (**Fig. 1D**). Autosomal and X chromosomal scHi-HOST sb-genes were tested for enrichment using a Fisher’s Exact Test, restricting analysis only to genes expressed in both LCLs and the tissue being compared. The greatest fold-enrichment was observed with the MAGE and GTEx LCLs. Whole blood and spleen, tissues with high lymphocyte counts, were also highly enriched for scHi-HOST sb-Genes. Interestingly, we additionally detected enrichment across nearly all tested tissues. Thus, sex biased differences in gene expression are highly reproducible across three LCL datasets and a subset is generalizable across tissues. In total, 849 (71.5%) of tested scHi-HOST sb-Genes were reproduced in at least one other dataset, with a core subset of 21 primarily X chromosomal genes previously identified to escape XCI^10^ showing sex-biased expression in all tested tissues. Interestingly, a single autosomal gene, *DDX43*, also showed conserved sex-biased expression in all tested tissues. This gene has previously been implicated in male fertility and spermatogenesis^22^, and displays sex-biased methylation in newborn ^23^ and adult ^24^ individuals.

## Partitioning of sb-Genes by mechanism

The variable effect size of sb-Genes points to different underlying mechanisms of sex-biased gene expression. The largest effects are due to the Y chromosomal genes that are entirely absent in females. We detected 14 Y chromosomal genes expressed in male LCLs. The next largest effect are genes that vary by copy number due to their presence on the X chromosome. In total, 92 X chromosomal genes display significant sex-biased expression, none of which lie in the pseudoautosomal regions shared by the X and Y chromosomes. Of these, 78 have previously been annotated as escaping X chromosome inactivation in at least one tissue^10^ and display a larger sex difference in expression (mean |log2FC|=0.38) than both sex-biased autosomal genes (mean |log2FC|=0.02) and sex-biased X chromosomal genes not annotated as XCI-escapees (n=14, mean |log2FC|=0.15). In total, 7.7% of sb-Genes can be explained due to a direct difference in copy number.

However, the vast majority of sb-Genes occur on the autosomes where they exhibit more modest effect sizes. One step upstream of these differences in gene expression is activation or repression by transcription factors (TFs), which have been previously shown to be involved in sex differences across mammalian lineages^25^. Thus, we hypothesized that some fraction of the remaining sb-Genes could be traced to variable expression of upstream TFs that lead to downstream targets exhibiting sex-biased expression. To determine if TF expression is predictive of sex-biased expression, we utilized a deep neural network (DNN) model previously trained and evaluated using >100,000 randomly drawn RNA-seq samples from the ARCHS4 resource^26^. This DNN predictor uses the combinatorial effects of TFs to predict the expression of all genes. We assigned TF expression in females as the baseline condition before applying the TF fold-changes determined from the differential expression analyses described above. Through this approach, we discovered that DNN-predicted target expression (based on 1600 TFs) agreed with our empirical dataset (Spearman’s ρ=0.25, p=1.72×10^-15^). We further hypothesized that only significantly sex-biased TF expression would be sufficient to predict genome-wide sex-biased expression. We thus restricted the DNN prediction to only include effects of sex-biased TFs (n=127, FDR<0.05) and discovered that DNN-predicted expression still correlated with our empirical data (Spearman’s ρ=0.20, p=4.53×10^-11^).

We then sought to identify and model the effects of the TFs that could be driving sex-biased expression. TF enrichment analyses through ChEA3 (which ranks likely causal TFs based on RNA-seq co-expression and ChIP-Seq datasets^27^) revealed 608 TFs with significant (FDR<0.05) enrichment for sex-biased targets (**Table S3**) in at least one dataset. Out of these enriched TFs, 81 display sex-biased expression themselves (FDR<0.05). By investigating the targets of enriched TFs, we discovered that the majority (79%) of sb-Genes are targets of at least one enriched sex-biased TF. An additional 10% of sb-Genes are targets of non-sex-biased TFs that were also enriched for sb-Gene targets. Thus, ∼89% of sb-Genes can be classified as possibly secondary to a difference in TF expression or activity (**Fig. 2A**).

**Figure 2.**
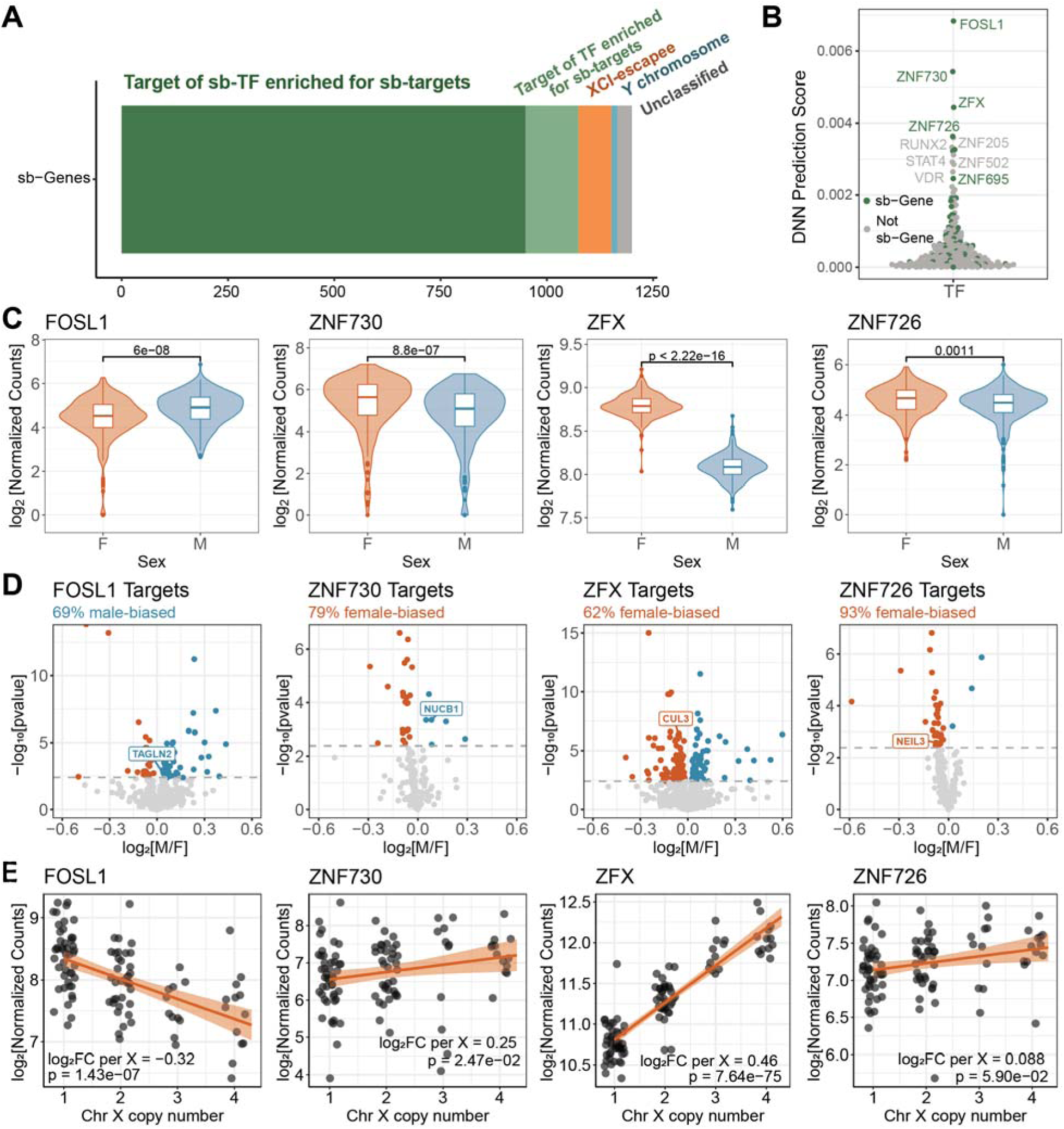
Nearly all sb-Genes can be explained by differences in sex chromosomes and their impact on sex-biased transcription factors. (**A**) Partitioning of sex-biased genes by location on the sex chromosomes (1.2% are on the Y; 6.5% are X-chromosome inactivation (XCI) escapees) and as enriched targets of transcription factors. (**B**) Ranking of ChEA3-enriched transcription factors by deep neural network (DNN) score where removal of a single transcription factor affects the correlation of the model. DNN score reflects the difference in Spearman correlation between predicted expression and true expression when either retaining all transcription factors or removing a single transcription factor from the model. Transcription factors are colored by sex-bias. (**C**) Differential expression of the top four enriched transcription factors by sex. P-value from Wilcox rank-sum test. (**D**) Volcano plot of scHi-HOST differential expression by sex results of all ChEA3 targets of each transcription factor. Orange and blue reflect significantly female- and male-biased targets respectively (FDR<0.05). (**E**) Expression of each transcription factor from LCLs with varying X chromosome copy number (data from^30^).

To further identify which TFs primarily contribute to sex-biased expression, we integrated enriched TFs with the DNN described above. By removing the effect of a single enriched TF at a time from the model, we were able to determine that the sb-Gene *FOSL1* has the largest impact on sex-biased expression (**Fig. 2B**). *FOSL1* encodes the Fos-like 1 (FOSL1, also called FRA1) protein which can act as both a transcriptional activator and repressor^28^. ScHi-HOST transcriptomics revealed that *FOSL1* displays significant male-biased expression (log2FC=0.37, p=7.39×10^-6^, **Fig. 2C**) and primarily male-biased sb-Gene targets (**Fig. 2D**).

Three other sb-TFs (*ZNF730, ZFX, ZNF726*) notably affected DNN model fit. In each case, we broadly see concordant direction of effect in the sex-biased expression of the TF and the sb-Gene targets (**Fig. 2C, D**). While little is known about *ZNF730* and *ZNF726*, *ZFX* is a highly female-biased gene that escapes X chromosome inactivation and has previously been shown to drive X-responsive expression across the genome^29,30^. Thus, for *ZFX*, the mechanism for female-biased TF expression is copy number variation. Previous work has demonstrated that much of the effect of additional copies of the X chromosome are mediated by ZFX and that a homologous TF encoded on the Y, ZFY, activates a similar set of targets, with ZFX having a larger effect^30^. In fact, *FOSL1*, *ZNF730*, and *ZNF726* all display at least nominal significance for correlation of expression with the number of X chromosomes in individuals with 1 to 4 X chromosomes (**Fig. 2E**; data from^29,30^). Notably for all 81 enriched sb-TFs that we identified, 49 (60.5%) are also correlated with X chromosome copy number (p<0.05; **Table S4**). Additionally, 29 sb-TFs (35.8%) are significantly differentially expressed in ZFX CRISPRi knockdown fibroblasts (p<0.05; **Table S4**), which supports previous work highlighting ZFX as a key transcriptional regulator of genome-wide sex differences^30^. However, the top autosomal sb-TFs previously discussed (*FOSL1, ZNF730, ZNF726*) are not significantly impacted during ZFX knockdown and display only limited correlation in expression (**Table S4**, **Fig. S2A**). This suggests that the sex-biased expression of most TFs that appear to mediate the sex-biased expression of autosomal genes may arise secondary to the number of X chromosomes, although this effect may be ZFX-independent.

Thus, through integration of ChIP-Seq signatures, coexpression datasets, and DNN modelling, we identified sb-TF expression as a likely mechanism for sex-biased expression across the genome and highlight four primary TFs driving these sex differences. Of these four, *ZFX* has higher expression in females due to females having two copies, while the sex-biased expression of the other three TFs can be traced back to X-chromosome copy number effects independent of ZFX.

## Modeling and testing the effect of a TF on sex-biased expression throughout the genome

We tested how well sb-Genes could be explained by the effects of sb-TFs through linear modeling and through examining experimental perturbation of sb-TF expression. First, the effects of each of the 81 enriched sb-TFs can be modeled based on two metrics from our dataset: 1) the relative expression in males vs. females for the TF (see Fig. 2C) and 2) the beta of the regression of TF on downstream target (**Fig. 3A**). Specifically, the expression level of the target gene can be modeled based on the relative increase in TF expression in one sex vs. the other (TF_sexeffect_) and the linear regression coefficient of the effect of TF expression level on target expression level within one sex (β) while accommodating the regression intercept (α) and residual (ε): *Target = α + β × (TF_measured_ × TF_sex_effect_) + ε.* The effect of sex-biased expression can then be simply modeled as a set of linear equations for all downstream targets of that TF.

**Figure 3.**
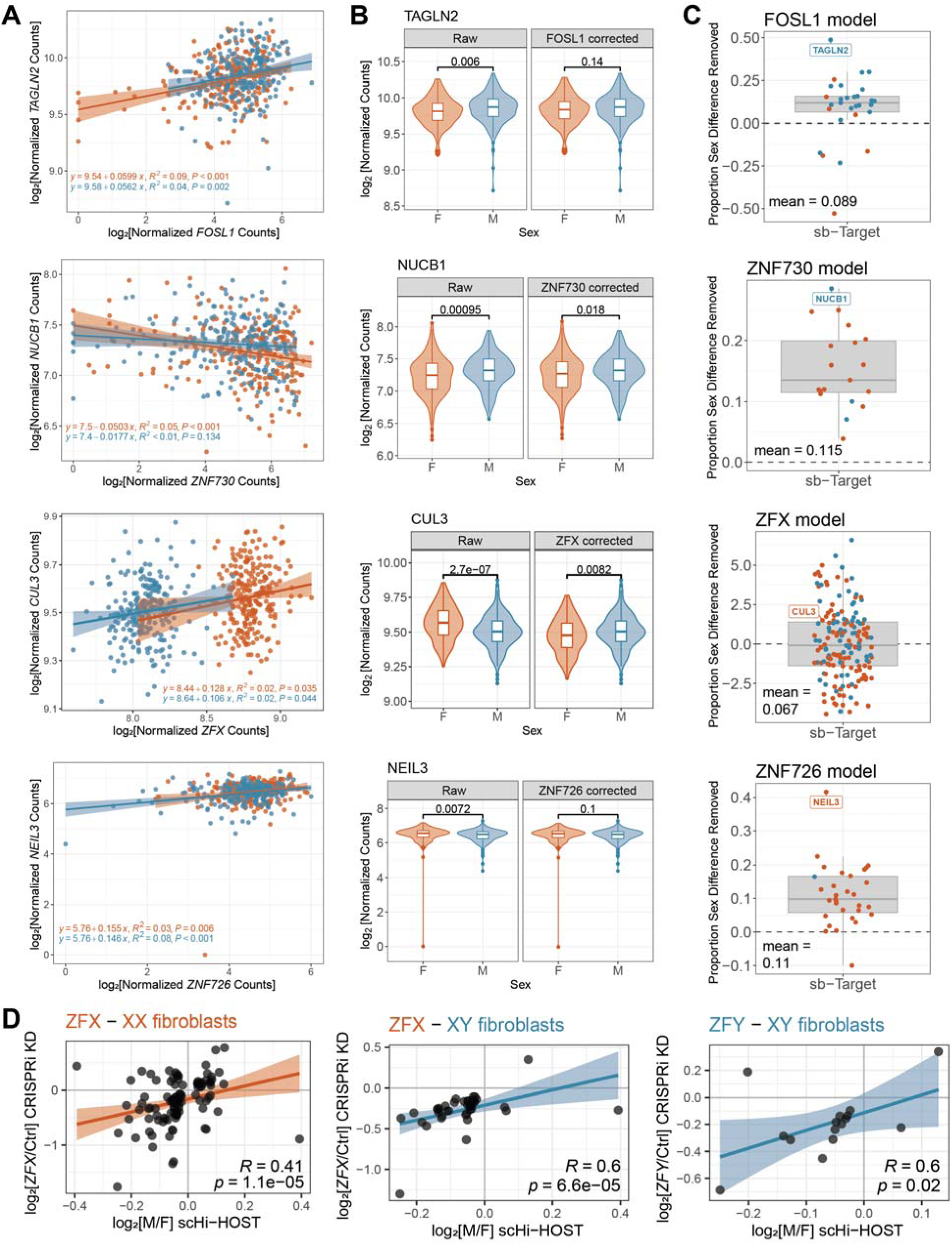
Transcription factor modeling of *FOSL1*, *ZNF730*, *ZFX*, and *ZNF726*. (**A**) Linear regression of TF expression and one of its sex-biased targets. Orange and blue denote female and male LCLs respectively. (**B**) Expression of a sex-biased target before and after linear model correction of TF sex-bias. Orange and blue denote female and male LCLs respectively. P-values from Wilcoxon Rank Sum test. (**C**) Linear modelling results per transcription factor where each target is plotted as a proportion of total sex difference removed by the model. Positive values indicate sex-difference removed through transcription factor modeling, whereas negative values indicate exacerbation of target sex-difference, with 1 representing complete remediation of target sex-difference. Orange and blue reflect significantly female- and male-biased targets respectively (FDR<0.05). (**D**) Correlation of scHi-HOST sex-biased expression and ZFX or ZFY CRISPRi knockdown differential expression of ZFX gene targets in XX of XY fibroblasts (data from ^2^). R and p-values from Spearman correlation.

We tested this model on the top four sb-TFs highlighted in the DNN models. We discovered that removing the sex difference in TF expression accounts for a mean of 11.8% of sex-biased expression in targets of FOSL1, ZNF730, and ZNF726 (**Fig. 3B, C**). Modeling results for the sb-TF ZFX revealed that ZFX sex-biased expression still accounted for 6.7% of target sex differences, but the model tended to over- or under-correct expression (**Fig. 3A, B, C**). This is likely due to the influence of ZFY, the Y chromosome homolog of ZFX.

We then tested the same models in the independent MAGE dataset. We again show that remediating the sex differential expression of *FOSL1* reduces the sex-bias in target expression (**Fig. S2B**), with parallel results also found for *ZNF730* and *ZNF726*.

Finally, for *ZFX* and its Y-chromosome homolog *ZFY*, RNA-seq has been published, defining how gene expression is affected by CRISPRi knockdown in fibroblasts^2^. We determined that targets of ZFX that display sex-biased expression in scHi-HOST are enriched in differentially expressed genes (FDR<0.1) during *ZFX* knockdown in both XX (fold enrichment=1.25, Fisher’s Exact Test p=0.001) and XY (fold enrichment=1.83, Fisher’s Exact Test p=0.0002) fibroblast cells. Similarly, we identified significant enrichment of sex-biased targets in differentially expressed genes (FDR<0.1) following *ZFY* knockdown in XY fibroblasts (fold enrichment=2.17, Fisher’s Exact Test p=0.004). We further identified a high degree of correlation between scHi-HOST differential expression directionality by sex and effects of *ZFX* knockdown in XX (Spearman’s ρ=0.41, p-1.1e-5) and XY (Spearman’s ρ=0.6, p-6.6e-5) cells as well as *ZFY* (Spearman’s ρ=0.6, p=0.02) knockdown (**Fig. 3D**).

Thus, our categorization of sb-Genes appears accurate following computational and experimental testing. Modeling the effects of sb-TFs based on our scHi-HOST dataset accurately predicts the expression of downstream target genes in two independent datasets and experimental knockdown supports these findings. The largest sex differences in gene expression are secondary to copy number variation, while most sex differences are found in autosomal genes and can be traced back to sex-biased TFs, the majority of which are influenced by copy number of the sex chromosomes.

## Identification of sb-eQTL with a two-step approach

While the large effects seen with sb-Genes on the X and Y are easily explained by their variation in copy number, and our evidence indicates that many of the smaller autosomal differences are secondary to these differences, some of these smaller differences might also be due to sb-eQTL. However, previous work has identified limited evidence of sb-eQTL through genome-wide interaction models^3,4^ or difference in slopes methods^5^. These methods support the discovery of large-effect sb-eQTL but are hampered by low power due to high multiple-testing burden. In addition, previous studies that have identified sb-eQTL have sampled populations with primarily European ancestry, further limiting discoveries of functional genetic variation. Therefore, we used a two-step pipeline in the diverse scHi-HOST cohort that reduces the multiple-test burden by first restricting analyses to conditionally independent eQTL in males or females separately before testing for a difference in slopes between sexes (**Fig. 4A**). We identified 5,502 conditionally independent eQTL in females and 5,740 eQTL in males in 4,599 and 4,718 eGenes (target genes of eQTL) respectively (q<0.05; **Table S5**). Interestingly, we identified an eQTL on the Y chromosome for the eGene TTTY14, demonstrating the first example of genome-wide significant eQTL on the human Y chromosome^6^ (**Fig. S3**). Combined, there were 10,529 unique eQTL for 6,331 eGenes, with 1,613 female-specific eGenes and 1,732 male-specific eGenes. We then tested all unique eQTL for a difference in slopes between males and females using the z-score method previously described^5,7^, and identified 2,390 sb-eQTL for 2,226 sb-eGenes (q<0.05), including 66 X-chromosomal genes (**Table S6**). The vast majority (90.2%) of discovered sb-eQTL display a significant effect in only one sex, although differences in amplification (7.4%) as well as rare instances of reverse effect between sexes (2.3%) are also evident (**Fig. 4B**). The majority of sb-eQTL are present in intronic regions, although several are also found in 3’ and 5’ UTRs and other annotated regulatory regions of the genome (**Fig. S3**).

**Figure 4.**
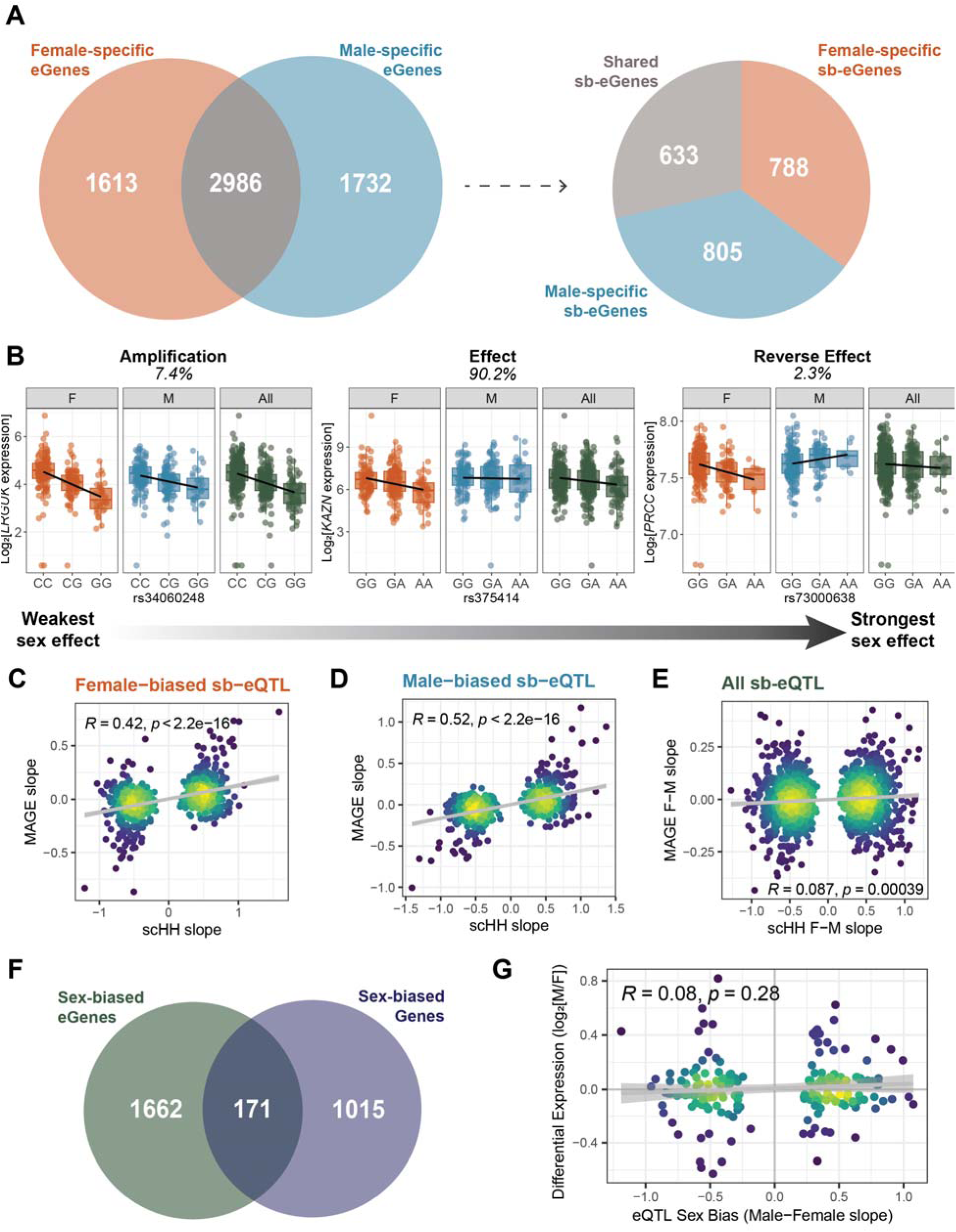
sb-eQTL are abundant throughout the genome but are of small effect size. (**A**) Discovery of sb-eQTL from conditionally independent eQTL through a two-step method reveals eGenes unique to each sex and variants with sex-biased association to nearby genes (**B**) Significant sb-eQTL classified into three main modes of action. (**C**) Replication of effect sizes of female-biased sb-eQTL in female MAGE data using Pearson correlation. (**D**) Replication of effect sizes of male-biased sb-eQTL in male MAGE data using Pearson correlation. (**E**) Replication of sex difference in effect sizes of sex-biased eQTL in MAGE data using Pearson correlation. (**F**). Overlap of sex-biased eGenes and sex-biased differentially expressed Genes displays no statistical enrichment (1.03-fold enriched; p=0.69). (**G**) Pearson correlation of eQTL sex-bias compared with differential expression effect from shared genes shows no significant correlation.

Through replication with an independent eQTL dataset (MAGE), we identified moderate replication at nominal p<0.05 of female-biased sb-eQTL (24.8%; **Fig. 4C**) and male-biased sb-eQTL (26.1%; **Fig. 4D**) with concordant effects in sex-stratified analyses. However, replication of sex-biased differences in slopes was minor (5.6% at nominal p=0.05), although we did identify significant correlation between scHi-HOST and MAGE effects (Pearson ρ=0.087, p=0.00039; **Fig. 4E**). We additionally conducted permutation analyses to identify possible biases in our two-step method. We found that permuting the sex prior to the first step of the pipeline led to similar numbers of sex-biased eQTL (**Fig. S3**), but that the concordance of effect with MAGE was only found with true sex (p<0.01; **Fig. S3**). Thus, although this two-step method revealed greater numbers of significant sb-eQTL than previous reports with significant concordance of effect in an independent dataset, we find that stratified approaches are prone to noise. Therefore, we conclude that at the current power, sb-eQTL cannot be detected at greater significance than observed by chance but that their direction of effect is more likely to be consistent across datasets.

Examination of the overlap between sb-Genes (genes with sex-biased expression; see Fig. 1) and sb-eGenes (genes regulated by sb-eQTL) demonstrated no enrichment (1.03-fold enrichment, p=0.69; **Fig. 4F**). For shared sb-Genes and sb-eGenes, we did observe slight correlation in effects, although this was also not significant (Pearson ρ=0.08, p=0.28; **Fig. 4G**) Thus, sb-eQTL do not appear to explain sex-biased expression, consistent with previous findings^1,5^.

## sb-eQTL underlie some sex-bias in human traits

Although most sex-biased gene expression is not due to sb-eQTL, we hypothesized that the cases where it does occur might underlie some of the sex differences seen in human phenotypes and disease risk. This is most pronounced in autoimmunity where diseases such as multiple sclerosis (MS) display severe sex-bias. To identify sb-eQTL associated with human disease risk, we utilized the genetic architecture tool iCPAGdb^8^. iCPAGdb integrates the NHGRI-EBI GWAS catalog^9^ with large datasets of plasma^10^ and urine^11^ metabolites and cellular host-pathogen phenotypes^12^ to determine shared genetic factors. Briefly, iCPAGdb overlaps lead SNPs from the integrated GWAS data with SNPs from a user-submitted list, accounts for linkage between signals, and calculates enrichment. We discovered that 173 phenotypes significantly overlap with our sb-eQTL (FDR<0.05, **Table S7**) with phenotypes such as body height and body mass index highly enriched (**Fig. S5**).

When results were restricted to disease susceptibility risk, we identified 47 significant enrichments with sex-biased autoimmune diseases predominantly enriched (FDR < 0.05, **Fig. 5A**). Due to the high enrichment of sb-eQTL in MS susceptibility loci (fold-enrichment=33.0, p= 2.89×10^-9^) and the relevance of lymphoblastoid cells to this disease (as EBV infection of B cells has been demonstrated to play a causal role^13,14^), we further investigated sb-eQTL effects in MS. Through integration of MS GWAS data from the International Multiple Sclerosis Consortium^15^, we identified 14 sb-eQTL (12 sb-eGenes) with genome-wide significant association (p<5×10^-8^) with MS, and an additional 4 (4 sb-eGenes) with suggestive association (p<1×10^-5^). We then tested all 18 associated variants for colocalization of GWAS signals with sex-stratified eQTL and identified 6 variants that show strong colocalization (PP4>0.7) in at least one sex (**Fig. 5B**). These eQTL results reflect sex-biased effects on known MS-risk genes such as CD58^16^, DDX39B^17^, GTF2H4^18^, and DEXI^19^.

**Figure 5.**
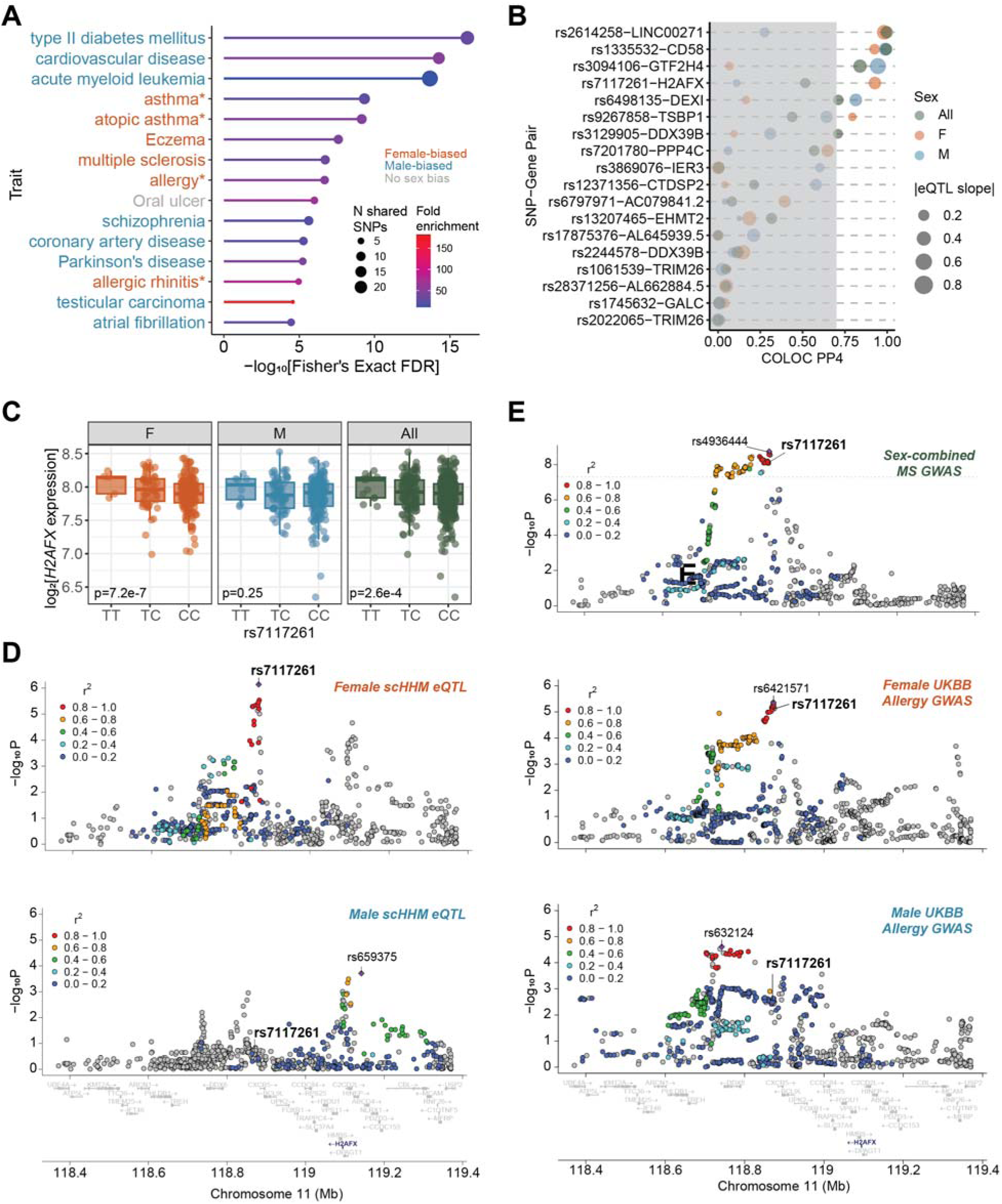
sb-eQTL are associated with disease-risk loci, particularly in autoimmunity. (**A**) iCPAGdb enrichment of sb-eQTL in human disease-risk loci. Colors denote sex-biased prevalence of each disease. * indicates interplay between sex and age where the disease is more common in adult females, but in ages pre-puberty are more common in males. (**B**) sb-eQTL colocalize with multiple sclerosis risk in a sex-dependent manner. PP4 > 0.7 signifies significant colocalization between eQTL and GWAS signals. Each line represents a unique SNP-gene pair with colors denoting PP4 values in female, male, and all individuals scHi-HOST eQTL analyses. Points are scaled by the absolute value of each respective eQTL slope to indicate effect sizes. (**C**) sb-eQTL rs7717261 displays greater association with *H2AFX* expression in females than in males. (**C**) LocusZoom plots of female and male eQTL nominal p-values from sex-stratified eQTL analyses. Colors denote linkage R^2^ value with index SNP (denoted by purple diamond) (**D**) LocusZoom plots of sex-combined multiple sclerosis GWAS nominal p-values and sex-stratified hayfever/allergy GWAS nominal p-values for the region surrounding *H2AFX*.

We identified a variant on chromosome 11 (rs7117261) that is associated with sex-biased expression of the nearby gene *H2AFX* (**Fig. 5C**), where there is only a significant association in female individuals (p=7.30×10^-7^; **Fig. 5D**) and not in males (p=0.25). The minor allele (T) is associated with increased risk of MS (**Fig. 5E**, p=7.16×10^-9^, OR=1.13). We then tested the sex-stratified eQTL and sex-combined MS GWAS for colocalization at this locus and found significant colocalization in female individuals (PP4=0.93) and not in male individuals (PP4=0.11) suggesting a sex-specific risk effect of *H2AFX* expression in MS pathogenesis. With sex-combined eQTL, we identified only moderate colocalization (PP4=0.52, **Fig. S4**), highlighting the importance of integrating sex-stratified analyses into genome-wide discoveries and suggesting that this effect is driven by the female eQTL association. Through a previously published phenome-wide association study (PheWAS) of UKBB sex-combined traits, we identified additional association of rs2614258 with doctor-diagnosed hayfever/allergy risk (p=1.6×10^-7^)^20^. Sex-stratified GWAS of the same phenotype revealed sex-bias in association (female p=8.0×10^-6^, male p=0.003). We again tested for colocalization, this time in each sex separately, and identified strong colocalization in female (PP4=0.94) eQTL, but not in male (PP4= 0.01) or sex-combined (PP4=0.56) analyses, mirroring the previous MS result (**Fig. 5E**, **Fig. S4**). *H2AFX* encodes the histone variant H2A.X. This histone variant becomes phosphorylated upon DNA double-strand breaks and has previously been primarily linked with multiple cancers including breast cancer^21^ and hepatocellular carcinoma^22^. However, previous work identified the upregulation of phosphorylated H2A.X in peripheral mononuclear blood cells (PBMCs) from patients with relapsing-remitting MS, the most prevalent form of the disease ^23^.

## Discussion

We identified sex-biased gene expression across the genome with 9% of genes displaying significant sex-bias. The largest sex differences in gene expression are the product of copy number variation due to the sex chromosomes, but these account for only 3% of sb-Genes in this study. However, the vast majority (79%) of sex-biased genes are targets of at least one sex-biased transcription factor, suggesting a possible mechanism of global sex differences in gene expression. One such enriched transcription factor*, ZFX*, is known to escape X chromosome inactivation and suggests that sex chromosome copy number is the underlying mechanism leading to sex-biased expression of *ZFX* targets. In fact, sex chromosome number may also underlie the sex-biased expression of autosomal TFs and drive sex-biased expression of targets. Thus, our data demonstrate that much of the observed sex-biased expression of human genes can be traced directly or indirectly to the number of X chromosomes, although our model cellular data represents cell-autonomous sex differences and does not account for the influence of sex hormones.

Beyond the effects of the X chromosome, the initial mechanism of sex-bias of autosomal genes may have additional contributing factors, such as differential chromatin accessibility between biological sexes^3^. In addition, a small portion of sex-biased genes (10%) are not targets of sex-biased transcription factors. These examples of sex-biased expression may be due to additive effects of multiple transcription factors or a possible amplification of minor sex differences in transcription factor expression. Lastly, not included in this study was the consideration of transcription-associated RNA-binding proteins or large-scale 3D-genome structure which may explain some residual sex differences in gene expression. But beyond categorizing sex-biased transcriptional effects, we have modeled the effects of the most important sex-biased TFs on their target genes and experimentally examined the effects of ZFX and ZFY in a loss-of-function transcriptomic dataset. In combining this categorization of sex-biased effects with modeling and empirical testing, we move to the level of molecular detail necessary to understand why individual genes have sex-biased expression and how this might be manipulated to impact human health. Future studies can further refine the impacts of FOSL1, ZNF730, and ZNF726 on sex-biased gene expression.

While it is well-accepted that sb-Genes are abundant throughout the genome, with many conserved across tissues^1^, the existence and relevance of sb-eQTL is much more contentious. By focusing on conditionally independent eQTL identified in each sex separately, we reduce the multiple-testing burden and have discovered extensive evidence of sex differences in *cis*-acting genetic regulation (2,390 sb-eQTL) that follow differences in amplification, effect in a single sex, and opposite effects in each sex. Although we are unable to find strong replication of sb-eQTL, consistent with previous reports^1,3,5^, we observed that the directionality of observed sb-eQTL was more strongly concordant in an independent dataset than with permuted data. This suggests there are true positive sb-eQTL in the dataset, but that with 480 LCLs, we are still underpowered and additional analyses with increased power are necessary to confirm small differences in sex-biased regulation. While not accounting for large-scale differences in sex-biased gene expression, sb-eQTL provide additional context for complex traits and diseases and may even reveal mechanisms that explain sex differences in disease risk and severity. Indeed, we identified sb-eQTL that show stronger association with human traits and better colocalization preferentially in one sex. In MS, a highly female-biased autoimmune disease, we identified sex-bias in known risk genes as well as discovered a novel gene connection (*H2AFX*) that also displays parallel sex-bias in susceptibility to hayfever and allergy. Further stratification of genome-wide discoveries by sex will continue to reveal novel biological mechanisms and support the inclusion of sex as a biological variable.

This work highlights the importance of investigating genomic data in a sex-stratified manner and is a call-to-action to continue to grow our efforts to improve the resolution of these small but widespread effects of sex across the genome.

## METHODS

### Lymphoblastoid cell lines

240 male and 240 female 1000 Genomes LCLs from 12 worldwide populations (Table S1) were purchased from the Coriell Institute. LCLs were maintained at 37°C in a 5% CO2 atmosphere and grown in RPMI 1640 media (Invitrogen) supplemented with 10% fetal bovine serum (FBS), 2 mM glutamine, 100 U/mL penicillin-G, and 100 mg/mL streptomycin. Equal numbers of pooled LCLs are stored in liquid nitrogen vapor phase.

### Single-cell transcriptomics

Equal numbers of LCLs were pooled and added to a 24-well non-tissue culture treated plate in PBS with 0.4% BSA, Mg^2+^, Ca^2+^, 100 U/mL penicillin-G, and 100 mg/mL streptomycin. Uninfected controls from the previously described infection^24^ were spiked after 3 h with 500 µL of the LCL growth media described above. After 24 h, pooled cells were collected, spun down, and resuspended in PBS with 0.04% BSA for single-cell cDNA library preparation. This process was repeated three times with 48 LCLs used in both scHi-HOST Neo (previously scHH-EIK in^24^) and scHi-HOST Morpheus (previously scHH-LGC in^24^) and 384 LCLs used in scHi-HOST Trinity (not previously described; Table S1).

Cell counts and viability were collected on a Guava EasyCyte HT system by 7-AAD staining before dilution to a solution of 1 million cells/mL with intended capture of 10,000 cells/well for scHi-HOST Neo and Morpheus and to 1.25 million cells/mL with intended capture of 20,000 cells/well for scHi-HOST Trinity. The 10x Chromium Single Cell 3’ platform version 3.1 (Pleasanton, CA) was used to generate individual barcoded cDNA libraries for each well following the manufacturer’s protocol. Briefly, the 10x Chromium Controller separates individual cells into nanoliter-scale gel beads in emulsion (GEMS) where cell-specific barcoding and oligo-dT-primed reverse-transcription occurs. For scHi-HOST Neo, 13,675 uninfected droplets were captured in 1 Chromium well. For scHi-HOST Morpheus, 36,716 uninfected droplets were captured across 2 Chromium wells. For scHi-HOST Trinity, 85,558 uninfected droplets were captured across 4 Chromium wells.

cDNA samples from scHi-HOST Neo were single-indexed and sequenced on an Illumina HiSeq system with a target depth of 50,000 reads per barcoded droplets. Reads were sequenced with read 1 length of 150 base pairs (bp) and read 2 length of 150 bp. This resulted in a mean depth per cell of 37,815 reads. scHi-HOST Morpheus samples were dual-indexed and sequenced on one Illumina NovaSeq S4 flow cell with target depth 100,000 reads per barcoded droplet. Reads were sequenced with read 1 length of 28 bp and read 2 length of 150 bp resulting in a mean depth per cell of 67,415 reads. Lastly, cDNA samples from scHi-HOST Trinity were dual-indexed and sequenced on a NovaSeq S4 flow cell with target depth of 50,000 reads per droplet. Reads were sequenced with read 1 length of 28 bp and read 2 length of 150 bp which resulted in a mean depth per cell of 69,933 reads.

### Single-cell RNA-seq alignment and LCL assignment

As previously described^24^, we processed all raw sequencing results using the 10X Genomics CellRanger 7.0.1 with default parameters unless otherwise indicated. Reads from each sample were mapped to GRCh38. For the single-indexed scHi-HOST Neo, we used the 10X Genomics Index-hopping-filter to removed index-hopped reads (https://github.com/10XGenomics/index_hopping_filter) before mapping to the human genome. To remove possible ambient RNA contamination, we used CellBender ^25^ v.0.3.0 with –fpr 0.01 and –epohcs 150. CellBender uses a deep generative model to learn a background noise profile to delineate cell-containing and empty droplets and provide noise-free gene count quantification.

We then used Demuxlet ^26^ to assign each barcoded read from cell-containing droplets to an LCL. Demuxlet utilizes a CellRanger bam file with barcoded sample reads and a VCF containing all autosomal genotypes (obtained from the 1000 Genomes Project, NYGC 30x high-coverage release) to computational deconvolute each droplet into its corresponding LCL identity. After assigning each droplet to its corresponding LCL, CellBender gene counts were pseudobulked by sum to produce gene counts for each LCL. To reduce any batch effects in expression between the scHi-HOST Neo, Morpheus, and Trinity datasets, we conducted ComBat-seq^27^ correction and merged all LCL gene counts into the combined dataset scHi-HOST Matrix which was used for all downstream analyses.

### Sex-biased differential expression

The R package “DESeq2”^28^ was then used to perform all differential expression analyses. Feature counts were normalized according to the DESeq2 default “Median of Ratios” method which accounts for differences in sequencing depth and RNA composition between samples. Genes with at least 5 normalized counts across 10% of individuals were retained for further analyses. Differentially expressed genes by sex were then discovered in DESeq2 with LCL population as an additional covariate to account for possible population effects. KEGG pathway enrichment was conducted using the R package “clusterProfiler”^29^.

A parallel analysis was conducted to discover sb-Genes in the MAGE dataset^30^. Raw counts were normalized as described above with LCL population as a covariate. Sb-Genes in GTEx were previously discovered using a linear model with covariates accounting for known sample and donor characteristics, as well as surrogate variables that capture hidden factors of expression variability^1^.

### Transcription factor enrichment and deep neural network modeling

Transcription factor enrichment of sb-DEGs was accomplished in ChEA3^31^ through the utilization of two co-expression datasets (ARCHS4^32^ and GTEx^33^) and two ChIP-Seq datasets (ENCODE^34^ and Literature^31^). Briefly, the overlap of sb-DEGs and known TF targets from these distinct datasets was compared and enrichment calculated with a Fisher’s Exact Test. Enriched TFs were then prioritized for additional analyses based on the Chea3 Mean Rank Score.

The deep neural network (DNN) model developed by Magnusson, et al. ^35^was used to predict gene expression from TF expression. This neural network was trained on the ARCHS4 expression database ^32^ using two hidden layers with 250 hidden nodes each. Using this model, we predicted sex-biased expression in scHi-HOST Matrix from TF expression and female/male fold-change results from the above DESeq2 analyses. We then compared the predicted values of sb-Genes to observed quantifications using Spearman’s correlation. To determine if sb-TFs are sufficient to predict sb-Gene expression, we retained only those TFs with significant sex-biased expression (n=127) and repeated the model as outlined above. Lastly, to prioritize TFs that highly impact sex-biased expression, we iteratively removed the effect of one TF at the time and evaluated the difference in model performance.

### Transcription factor linear modeling and enrichment in knockdown datasets

We utilized linear models to determine the effect of removing the observed sex-bias in TF expression on target genes. We tested all sb-Genes annotated as targets of a particular TF in Chea3 for co-expression by generating a linear model of target expression as a function of TF expression. Target and TF expression matrices were generated as outlined above in the DESeq2 differential expression analyses. We then modeled how the removal of sex differential TF expression would impact expression of those targets with significant (p<0.05) co-expression. We utilized a model based on the relative increase in TF expression in one sex vs. the other (TF_sexeffect_) and the linear regression coefficient of the effect of TF expression level on target expression level within one sex (β) while accommodating the regression intercept (α) and residual (ε): *Target = α + β × (TF_measured_ × TF_sex_effect_) + ε*. To determine which targets were most impacted by sb-TF expression, we compared the modulated log2(M/F) fold changes of each target to the baseline values. For *ZFX* CRISPRi knockdown, we utilized DESeq2 differential expression results previously published in^2^.

### Sex-biased eQTL discovery

Sex-biased eQTL were discovered through a two-step method executed primarily in tensorQTL^36^ in Python v.3.11.4. First, scHi-HOST Matrix counts in each sex separately and sex-combined were normalized through the median of ratios method in DESeq2. Genes with at least 6 unnormalized counts across 20% of individuals were retained for eQTL discovery. Gene counts were then further normalized using rank-based inverse normal transformation as previously described^37^. To account for population structure, we utilized the top 5 genotypic principal components (PCs) from the autosomal genome as covariates. In addition, the top 15 probabilistic estimation of expression residuals (PEER) factors^38^ were used to account for hidden confounding variables in the expression matrix. PEER factors were tested for collinearity with known covariates such as known genotypic PCs and sex (sex-combined analysis only). The adaptive permutation mode (--permute 1000 10000) of tensorQTL was conducted in each sex with the covariates described above in a 1 Mb window from the transcriptional start site (TSS) of each gene to account for multiple tests within each gene. Multiple test correction across the genome was then conducted using the Storey’s Q^39^ method on the top eQTL for each gene to determine a significance threshold (q<0.05) for each gene. Conditionally-independent *cis*-eQTL were then discovered in each sex using an iterative linear regression model executed in tensorQTL with the significance threshold of q<0.05 for each gene.

We then tested all unique conditionally independent *cis-*eQTL for a difference in slopes between females and males with the following z-test method previously described^7^:

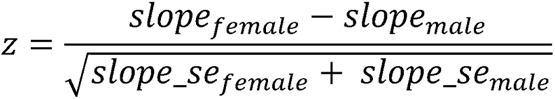

Resulting p-values were then corrected using the Storey Q method (q<0.05) to determine significant sb-eQTL.

Discovered sb-eQTL were classified into amplification, effect in a single sex, and reverse effect. The classification for amplification was if a sb-eQTL was genome-wide significant in one sex, nominal p<0.05 in the other sex, and the slopes are concordant between both sexes. Reverse effect was categorized as genome-wide significant in one sex, nominal p<0.05 in the other sex, and the slopes are discordant between both sexes. Effect in a single sex was categorized as any sb-eQTL that was genome-wide significant in one sex, with nominal p>0.05 in the other sex.

### Phenotype association of sex-biased eQTL

To investigate possible phenotypic associations of sb-eQTL, we utilized the command-line execution of the interactive Cross-Phenotype Analysis of GWAS database (iCPAGdb^8^). Briefly, iCPAGdb trims an input list of SNPs (here sb-eQTL) to leading variants based on linkage disequilibrium information from 1000 Genomes European populations^40^. These variants were then queried against cataloged GWAS variants from the NHGRI-EBI GWAS catalog^9^, urine ^11^ and plasma metabolites^10^, and Hi-HOST infectious disease phenotypes^12^. Both direct overlap and overlap with SNPs in linkage disequilibrium (r^2^>0.4) with lead variants were considered. Enrichment of query SNPs within GWAS phenotypes was calculated based on observed vs. expected overlap and significance calculated with Fisher’s Exact Test. Phenotypes with an enrichment FDR<0.05 were considered to share significant genetic architecture with sb-eQTL. Phenotypes were filtered to include only single effect phenotypes and behavioral GWAS were removed due to their likelihood to be biased by misreporting^41^.

### Colocalization of phenotype-associated sex-biased eQTL

We applied Giambartolomei et al.’s colocalization analysis (COLOC), using the R package “coloc”^42^ to determine if GWAS and eQTL signals were due to the same causal SNP. COLOC uses a Bayesian framework to calculate the posterior probabilities that two traits are not associated in the locus of interest (PP0), only one trait is associated in the locus (PP1 and PP2), both traits are associated at the locus but with different, independent causal variants (PP3), or both traits are associated with a single causal variant in the locus (PP4). For GWAS summary statistics, we filtered SNPs within a 100 kilobase (kb) window from the SNP of interest. For eQTL datasets, we filtered all eQTL for a candidate eGene and then filtered SNPs within a 100 kb window from the SNP of interest. We ran the COLOC “coloc.abf” function using the default prior parameters, p1=1×10^-4^, p2=1×10^-4^, and p12=1×10^-5^ for all analyses. PP4 between 0.700 to 0.900 was interpreted as likely to share a single causal variant, while PP4>0.900 was interpreted as sharing a single causal variant. The PP4/PP3 measured the intensity of the colocalization signal with values >5.00 indicating further support for colocalization and >3.00 suggesting likely colocalization^43,44^. LocusZoom plots were created using “locuszoomr”^45^.

## Data Availability

All data produced in the present study are available upon reasonable request to the authors

## Acknowledgments

We thank the investigators and individuals from diverse populations genotyped as part of the 1000 Genomes Project who have made their LCLs available through the Coriell Institute. We thank the individuals who participated in the UK Biobank and International Multiple Sclerosis Genetics Consortium (IMSGC) studies as well as the investigators for helpful distribution of data. We thank members of the Ko lab for useful discussion. We acknowledge the assistance of the Duke Molecular Physiology Institute Molecular Genomics Core for the generation of data for the manuscript. We thank Nikolai G. Vetr and Craig Lowe for helpful discussion. AGJ, GC, LW, BS, and DCK were supported by NIH grant R01AI170089. TD, LW, and DCK were supported by NIH grant R01AI118903. LW, BS, and DCK were supported by R21AI153812. AGJ, BS, and GC were supported by Duke Precision Genomics Center student pilot grants. AGJ and TD were supported by TriCEM Graduate Student Fellowships. ASKR was supported by the Whitehead Scholar Award.

## Author contributions

A.G.J. and D.C.K. designed the study. B.S. and G.G.C conducted sample preparation. GWAS phenotype enrichment and colocalization studies were completed by A.G.J., L.W., and T.D. A.K.S.R, T.D., and A.G.J. conducted data visualization. All other analyses were completed by A.G.J. Manuscript written by A.G.J. and D.C.K. with input from all co-authors.

## Data and code availability

eQTL summary statistics and differential expression results and code for both scHi-HOST and MAGE analyses will be available at Mendeley Data as of the date of publication (DOI: 10.17632/7mhjbwktvg.1). Individual LCL genotyping data is available through 1000 Genomes (FTP: http://ftp.1000genomes.ebi.ac.uk/vol1/ftp/). Single-cell RNA-seq data for scHi-HOST Neo and Morpheus are available from GEO (GSE205796). Single-cell RNA-seq data from scHi-HOST Trinity will be available from GEO as of the date of publication. All other data are available in the main text or the supplementary materials.

## Supplementary information

Document S1. Figures S1-S5

Table S1. Excel file containing 1000 Genomes LCL sample table from scHi-HOST Matrix. Related to Figure 1.

Table S2. Excel file containing scHi-HOST Matrix differential gene expression by sex, DESeq2 results. Related to Figure 1.

Table S3. Excel file containing ChEA3 significantly enriched transcription factors for scHi-HOST Matrix sb-Gene targets (FDR<0.05). Related to Figure 2.

Table S4. Excel file containing ChEA3-enriched sex-biased transcription factors with expression change per copy of Chr X or Chr Y in LCLs and effect of ZFX CRISPRi knockdown in fibroblasts. Related to Figure 2.

Table S5. Excel file containing scHi-HOST Matrix conditionally-independent sex-stratified eQTL tensorQTL results (q<0.05). Related to Figure 4.

Table S6. Excel file containing scHi-HOST Matrix significant sb-eQTL tensorQTL results (q<0.05). Related to Figure 4.

Table S7. Excel file containing iCPAGdb significantly enriched phenotypes for scHi-HOST Matrix sb-eQTL (q<0.05). Related to Figure 5.

