## Supplemental Figures for "Biological sex affects gene expression and functional variation across the human genome"

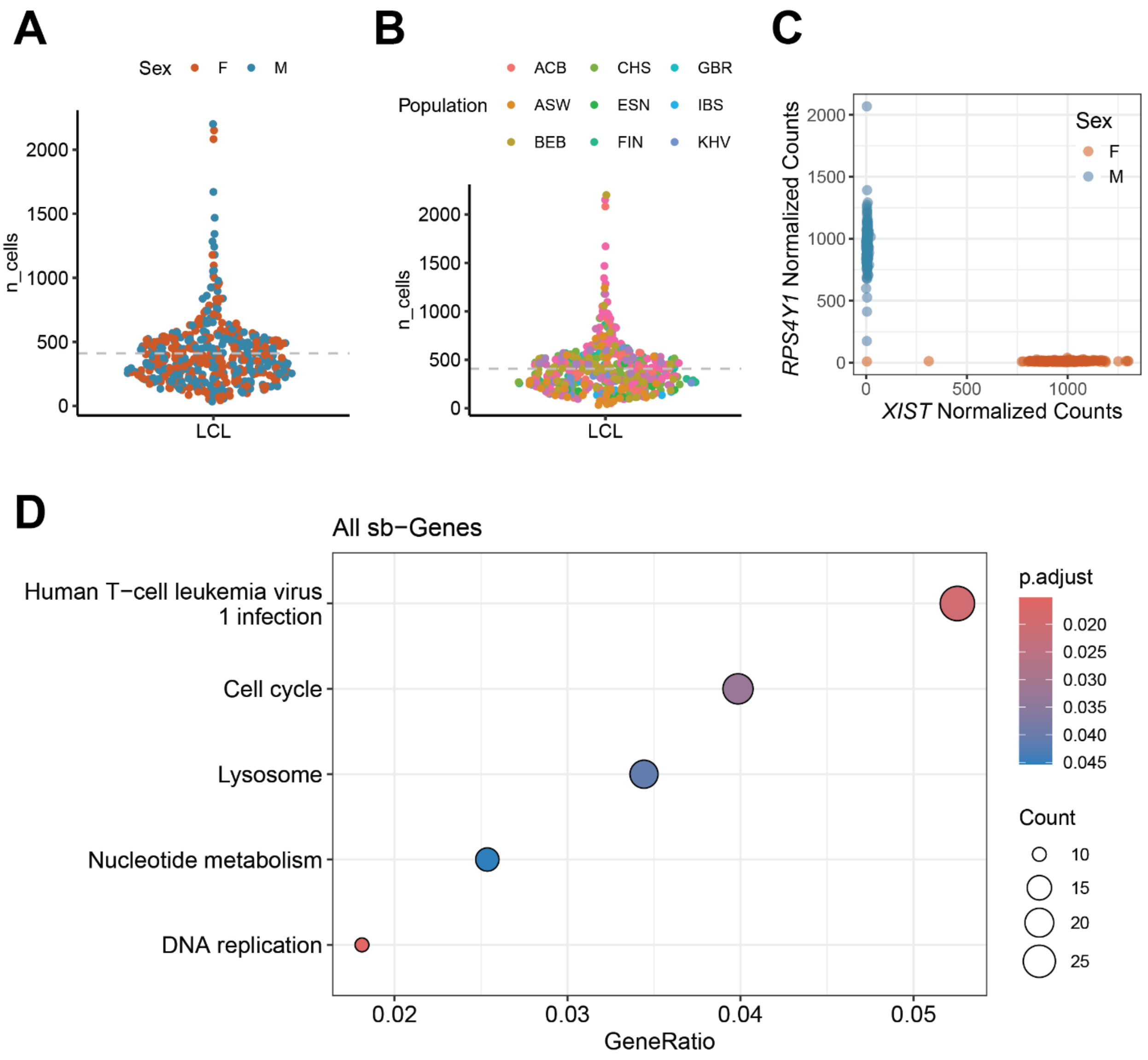


**Fig. S1.**

scHi-HOST Matrix sequencing and differential expression. Number of recovered singlets assigned to individual LCLs by (**A**) sex and (**B**) population. (**C**) Expression of Y chromosomal gene *RPS4Y1* and XCI-escaping gene *XIST* in scH-HOST Matrix LCLs. (**D**) KEGG pathway enrichment of scHi-HOST Matrix sb-Genes.

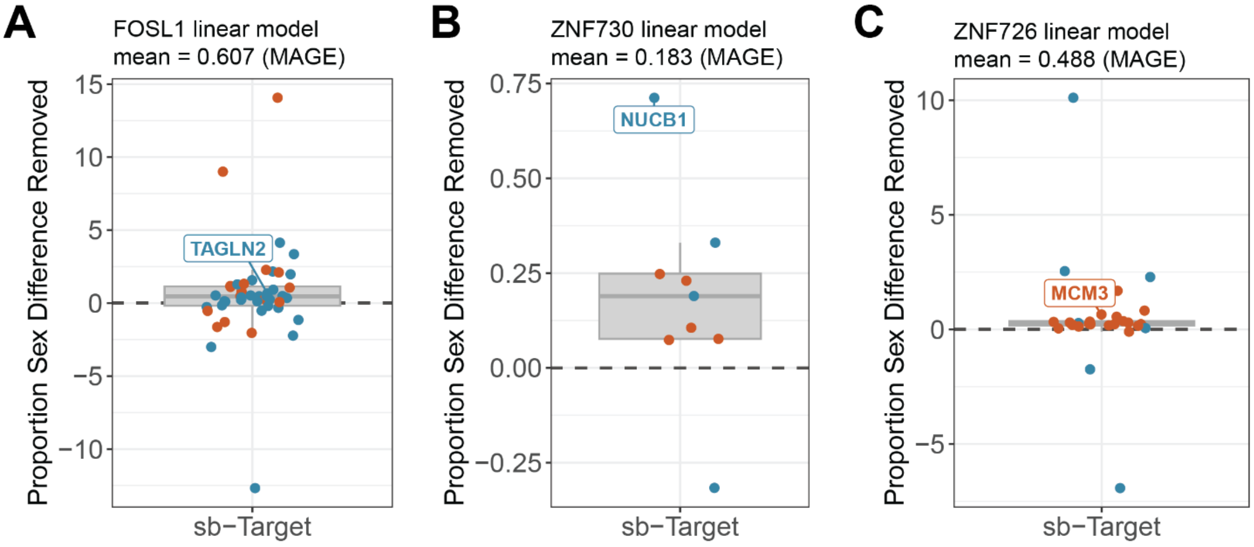


**Fig. S2.**

Linear modelling results in MAGE expression data for (**A**) *FOSL1*, (**B**) *ZNF730*, and (**C**) *ZNF726* where each target is plotted as a proportion of total sex difference removed by the model. Orange and blue reflect significantly female- and male-biased targets respectively (FDR<0.05).

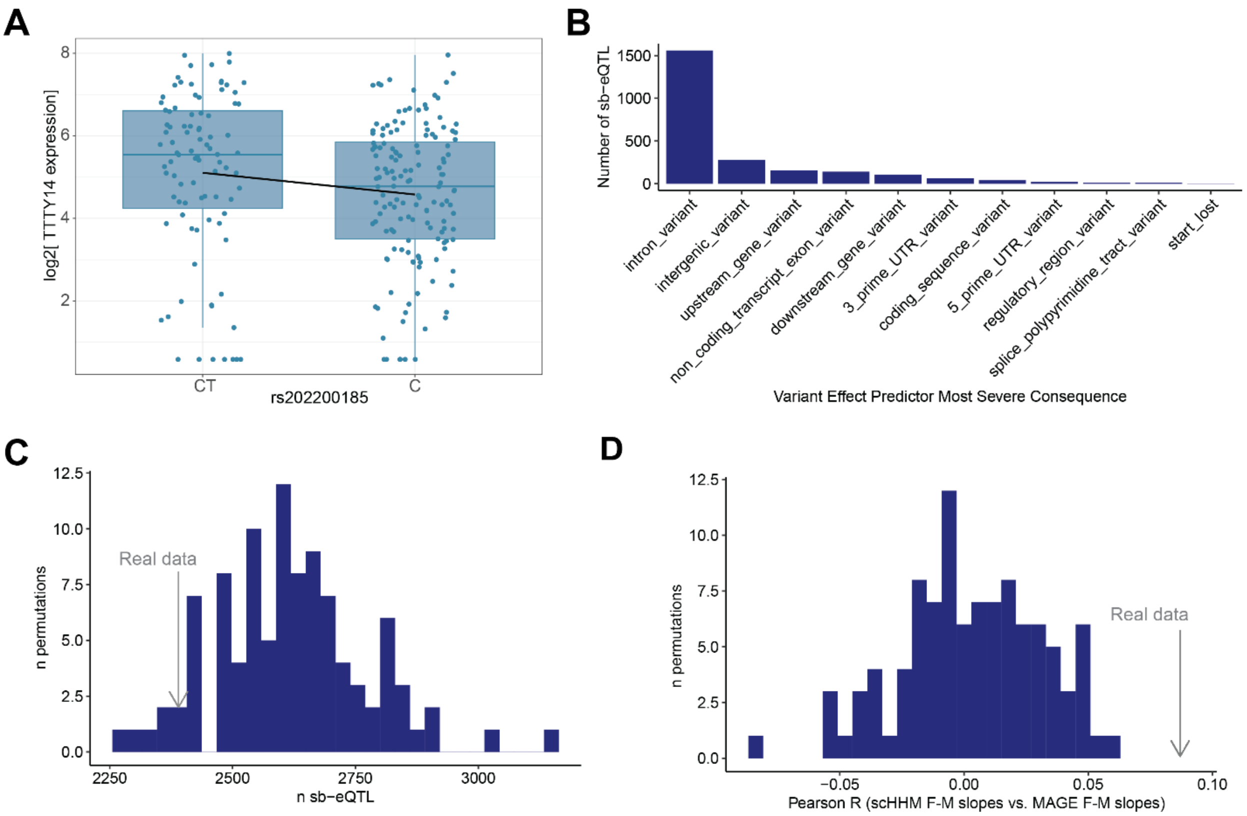


**Fig. S3.**

Sb-eQTL in scHi-HOST Matrix. (**A**) rs202200185 is an eQTL for the Y-chromosomal gene TTTY14. (**B**) sb-eQTL plotted by Ensembl Variant Effect Predictor most severe consequence. (**C**) Distribution of 100 scHi-HOST permutations and number of significant interaction eQTL ($q<0.05$). (**D**) Distribution of 100 scHi-HOST permutations by correlation of sex-biased effect (difference between female and male slopes for each sb-eQTL) with true sex MAGE results.

**A**

| eQTL Sex | Trait | PP0 | PP1 | PP2 | PP3 | PP4 |
|---|---|---|---|---|---|---|
| Female | MS | 4.08e-06 | 0.0267 | 7.17e-06 | 0.0461 | 0.927 |
| Male | MS | 8.67e-05 | 0.837 | 5.11e-06 | 0.0493 | 0.113 |
| All | MS | 7.30e-05 | 0.450 | 5.79e-06 | 0.0351 | 0.515 |
| Female | Hayfever/allergy female | 0.00567 | 0.0168 | 0.0101 | 0.0289 | 0.939 |
| Male | Hayfever/allergy male | 0.788 | 0.166 | 0.0263 | 0.00553 | 0.01450 |
| All | Hayfever/allergy all | 0.00251 | 0.399 | 0.000218 | 0.034 | 0.564 |

**B**

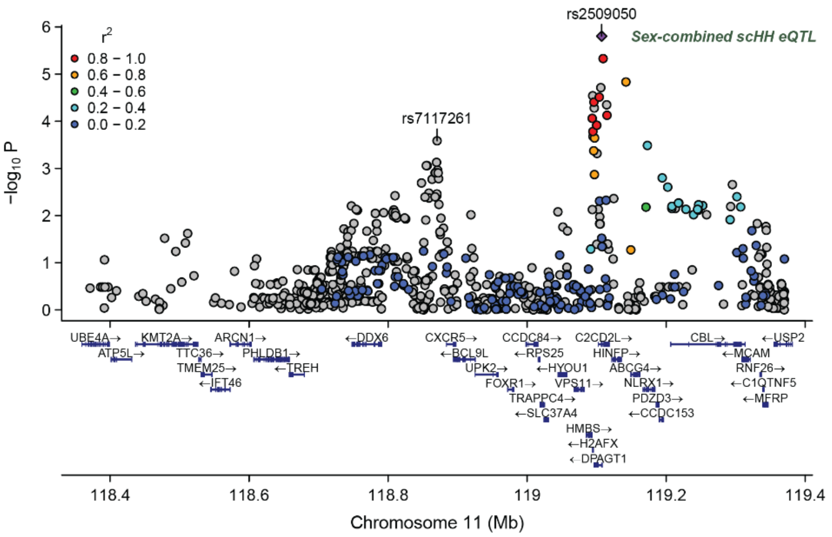


**C**

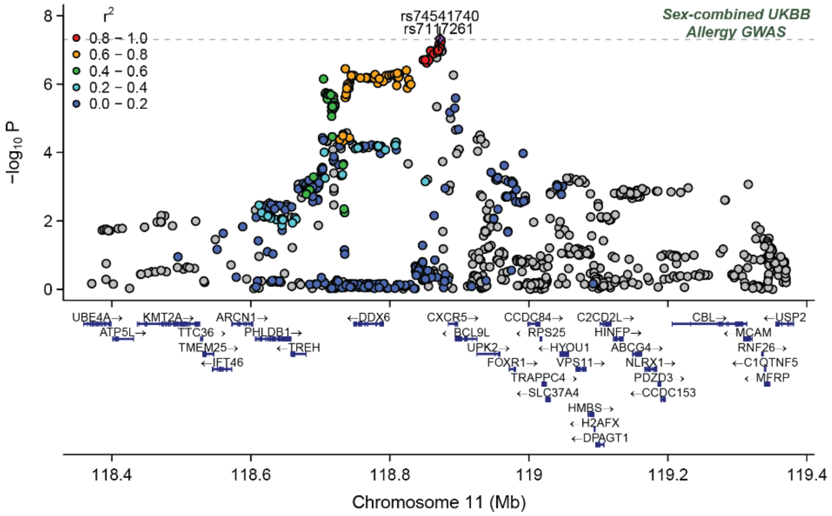


**Fig. S4.**

Sb-eQTL overlap GWAS phenotypes. (**A**) COLOC statistics for rs7117261-H2AFX associations with multiple sclerosis and UKBB hayfever/allergy GWAS. (**B**) LocusZoom plots of sb-eQTL rs7117261 in sex-combined scHH Matrix eQTL *H2AFX* association. (**C**) LocusZoom plot of sb-eQTL rs711726 displays association in sex-combined UKBB hayfever/allergy GWAS.

**Table S1. (separate file)**

1000 Genomes LCL sample table from scHi-HOST Matrix. Related to Figure 1.

**Table S2. (separate file)**

ScHi-HOST Matrix differential gene expression by sex, DESeq2 results. Related to Figure 1.

**Table S3. (separate file)**

ChEA3 significantly enriched transcription factors for scHi-HOST Matrix sb-Gene targets (FDR<0.05). Related to Figure 2.

**Table S4. (separate file)**

ChEA3-enriched sex-biased transcription factors with expression change per copy of Chr X or Chr Y in LCLs and effect of ZFX CRISPRi knockdown in fibroblasts. Related to Figure 2.

**Table S5. (separate file)**

ScHi-HOST Matrix significant conditionally-independent sex-stratified eQTL tensorQTL results (q<0.05). Related to Figure 4.

**Table S6. (separate file)**

ScHi-HOST Matrix significant sb-eQTL tensorQTL results (q<0.05). Related to Figure 4.

**Table S7. (separate file)**

iCPAGdb significantly enriched phenotypes for scHi-HOST Matrix sb-eQTL (FDR<0.05). Related to Figures 4 and 5.
